# Exploring Views of Technology to Inform Maternal and Child Health Interventions for Black Birthing People

**DOI:** 10.1101/2022.11.28.22282786

**Authors:** Elisha Jaeke, Jessica Olson, Dalvery Blackwell, Sophia Aboagye, Reiauna Taylor, Kaija L Zusevics, Melinda Stolley

## Abstract

**Background:** Black birthing people experience disproportionately higher rates of morbidity and mortality, and poorer infant health outcomes than all other racial and ethnic groups. Statewide in Wisconsin, an alarming disparity exists between Black and Non-Hispanic White (NHW) breastfeeding initiation rates (54% and 87%, respectively). For the last fourteen years, the African American Breastfeeding Network (AABN) has operated in Milwaukee County, where 70% of the state’s Black population lives. AABN’s mission is to improve maternal health and champion breastfeeding equity for Black birthing families. In 2018, AABN received more referrals than could be served by their team of doulas, lactation consultants, and advocates, leading our team to pursue ways to expand their reach through technology-based approaches. With the onset of COVID-19, the need for telehealth support increased drastically, as referrals for AABN services tripled. In this study, we explore the use of technology to support Black birthing families in Wisconsin and beyond, in the hopes of expanding the capacity of organizations like AABN.

**Methods:** A survey was conducted amongst 21 Black women who were pregnant or had given birth in the last year, regarding perceptions of technology. Women were split equally into three focus groups to discuss survey results further and provide context. Focus groups were recorded, transcribed, and coded for themes.

**Results:** Participants were “Likely” to use their phone or the internet to look for general or health-related information, but only “Somewhat Likely” to trust either general or health-related information. Participants were “Somewhat Likely” to trust the information on the phone or the internet as much as their healthcare provider (doctor, nurse, doula) in person. In the focus groups, all participants agreed that the phone or the internet was the first place looked to for information, ahead of consulting a healthcare professional. A comprehensive list of desired features was generated to guide the development of a resource to support AABN.

**Conclusions:** Study findings support exploring mobile technology-based approaches for breastfeeding support and education. Participants emphasized that to be relevant for them, a telehealth resource would need to: 1) Build meaningful connections between patients and providers, 2) Treat diverse opinions with trust and respect, and 3) Offer multiple sources and formats for information that can be easily cross-referenced.

## INTRODUCTION

Black birthing families experience disparate health outcomes compared to their white counterparts. Of the 371 infant deaths in Wisconsin in 2019, 23.45% (n=87) were African American infants, even though AA infants made up only 11.21% of total births that year^1^ Nationally, in 2019, infant mortality rates were 2.4 times higher in AA births compared to non-Hispanic Whites (NHW) (10.62 vs. 4.49 per 1,000 births, respectively).^2^ Leading causes of infant mortality were low birthweight, congenital malformations, maternal complications, accidents, and sudden infant death syndrome.^3^

In 2008, the African American Breastfeeding Network (AABN) was established to improve maternal and child health and breastfeeding equity with community-based, family-centered, culturally tailored support.^4^ AABN advocates for system- and policy-level changes, partners with community allies, and implements health education, lactation, and doula support services. AABN has helped over 4,000 Black families achieve their birth experience and breastfeeding goals, building strong communities with increased capacity to support breastfeeding access and reduce stigma. AABN uses evidence-based approaches to improve maternal health outcomes. Doula-assisted mothers are four times less likely to have babies with low birth weights, two times less likely to experience a birth complication involving themselves or the baby, and significantly more likely to initiate breastfeeding.^5^ Long-term, breastfeeding is protective against breast cancer, ovarian cancer, and type 2 diabetes.^6^

Before the coronavirus disease 2019 (COVID-19) pandemic, there was interest in gathering Black birthing people to understand how technology might expand some of the successes of AABN programming beyond its geographic region and capacity. However, with the onset of the COVID-19 pandemic, additional challenges were presented to the current maternal and infant health crisis. Referrals to AABN’s services tripled, due to many public health resources shifting their focus to the immediate needs of the pandemic, and away from maternal and infant health.

COVID-19 forced many health services to shift online. By April 2020, telemedicine utilization surged to 46%, compared to 11% pre-pandemic.^7^ As the pandemic disparately impacted vulnerable communities, the rapid change to healthcare access exacerbated an existing digital divide. Black patients exhibited lower levels of telehealth utilization than NHW counterparts and were more likely to continue to seek in-person urgent care.^8^ Additionally, patients who were Black, female, and/or of lower socioeconomic status were less likely to complete scheduled telehealth visits.^9^ Disparities caused by structural racism and other social determinants of health affect one’s access to trusted healthcare providers and quality healthcare. Across pregnant populations of all races and ethnicities, severe increases in maternal mental health issues and domestic violence occurred, while prenatal visits decreased.^10^ When virtual healthcare services fail to address these disparities, gaps widen.

In this study, we show how data initially collected to inform the dissemination of best practices used by AABN was used to build telehealth resources in response to COVID-19. By understanding engaging formats, key features, and factors that inspire trust in technology, our data-driven approach resulted in the launch of a successful telehealth-based intervention to support Black birthing families in Wisconsin.

## METHODS

### Recruitment

In the summer of 2018, Black birthing people were recruited by AABN staff, based on past referrals to AABN services. 25 women were invited to participate, and 21 were accepted (Table 1). Inclusion criteria were: 1) over 18, 2) self-identify as African American/Black, 3) able to read, and speak English, and 4) either currently pregnant or have given birth in the past year. No restrictions were made on gender, but no participants self-identified as non-binary, gender non-conforming, or men.

**Table 1.** Demographic Characteristics of Participants

| Table 1. Demographic Characteristics of Participants |  |  |
| --- | --- | --- |
|  | n | % |
| Race |  |  |
| · African American/Black | 21 | 100% |
| Gender |  |  |
| · Woman | 21 | 100% |
| Age |  |  |
| · 18-25 | 6 | 29% |
| · 26-35 | 10 | 47% |
| · 36-45 | 5 | 24% |
| Highest Level of Education Completed |  |  |
| · High School or Equivalent | 6 | 29% |
| · Technical/Occupational Certificate | 5 | 24% |
| · Associate Degree | 4 | 19% |
| · Bachelor's Degree | 4 | 19% |
| · Post-Baccalaureate Degree | 2 | 9% |
| Total Annual Household Income |  |  |
| · < \$25,000 | 7 | 33% |
| · \$25,000 – \$50,000 | 6 | 29% |
| · \$50,000 - \$100,000 | 6 | 29% |
| · \$100,000 + | 2 | 9% |

### Study Setting

All surveys and the three focus groups were conducted on the same day in a community setting with ample parking, refreshments, and free childcare. Participants were asked to check in and verify the accuracy of the inclusion criteria. Each participant chose a unique identifier and filled out survey questions on iPads. At the end of the survey and focus group, participants received $50 for their time and willingness to share perspectives.

### Survey Format and Analysis

Survey questions were asked on electronic tablets using Poll Everywhere software (Poll Everywhere, Inc. San Francisco, CA, USA). Women signed in using their real names to receive the incentive for participation, but survey responses were kept anonymous with the unique identifier privately selected by the participants. Participants were asked to complete a survey on electronic tablets provided by the facilitators. Participants entered a randomly chosen identifier and completed the survey questions anonymously (Table 2). Questions had the options “Very Unlikely”, “Unlikely”, “Somewhat Likely”, “Likely”, and “Very Likely”, numerically coded as 1-5, respectively. Responses were reported as averages with standard deviation.

**Table 2.** Survey responses from Black birthing women regarding perceptions of technology.

|  |  |
| --- | --- |
|  | Average ± SD |
| How likely is it that you will use your phone/internet to look for answers to questions? | 4.20 ± 1.06 |
| How likely is it that you will use your phone/internet to look for answers to health-related questions? | 4.23 ± 1.00 |
| How likely are you to trust information found on your phone/internet? | 3.33 ± 0.91 |
| How likely are you to trust information about your health that you find on your phone/internet? | 3.04 ± 0.80 |
| How likely is it that you will use new technology if it is free? | 4.19 ± 0.98 |
| If presented in a trustworthy way, how likely are you to trust information you find on your phone/internet as much as information from a health professional (doctor, nurse, doula) in person? | 3.62 ± 1.07 |

### Focus Group Format and Analysis

After survey completion, aggregated results were projected on a screen and used to guide focus group questions. Two facilitators led the focus groups with a trained assistant as a note-taker. Facilitators paid attention to the voices in the room and encouraged those who had not spoken to weigh in before moving to the next question, to help ensure that all voices were heard. The research team was entirely comprised of women, and 3 of the 4 research team members present self-identified as African American. Focus groups each lasted about 90 minutes and were digitally recorded and transcribed. Transcripts were compiled using ATLAS.ti Qualitative Data Management Software (ATLAS.ti, Berlin, DE, EUR) for analysis. Research team members coded each of the three focus group transcripts guided by a team-developed codebook looking for positive comments, negative comments, and features desired in technology designed to improve maternal and child health. After independent coding by each team member, the entire research team discussed the content to come to a consensus.

### Data Validation

The research team returned to present their findings in a town hall setting with all study participants. Participants had an opportunity to immediately provide feedback and were supplied with slides and contact information to review conclusions independently and contact the team with corrections.

### Ethical Compliance

All data collection and research-related actions were completed under approval from the academic medical center’s Institutional Review Board (PRO#00032076)

## RESULTS

### Survey Results

Participants were “Likely” to use their phone or the internet to look for general or health-related information, but only “Somewhat Likely” to trust either general or health-related information. Participants were “Somewhat Likely” to trust the information on the phone or the internet as much as their healthcare provider (doctor, nurse, doula) in person. Participants were likely to use new technology if it was provided free of charge.

### Thematic Results from Focus Groups

All focus groups agreed that the phone or the internet was the first place looked to for information, ahead of consulting a health professional. While participants endorsed technology as a fascinating and necessary way to stay connected and acquire knowledge, it was also shared that technology is intimidating and requires caution when examining sources. The ability to cross-reference and validate information was highly valued amongst focus group participants.

When considering digital delivery of healthcare services, a format like WebMD with easy provider access and symptom-checking software was suggested. The incorporation of homeopathic treatments in addition to allopathic medicines was endorsed. Visual tutorials were recommended to educate and provide clients with tools for success. Connection points were another desire of any digital programming technology, and participants expressed interest in message boards that could connect individuals with fellow community members with similar experiences.

For health services utilized remotely, participants underscored the importance of culturally relevant advice, as well as the ability to give and receive honest, realistic feedback. Participants also sought the ability to connect virtually with a consistent professional through FaceTime or a similar platform, especially during pregnancy and postpartum periods. Additionally, it was agreed upon that it is necessary to embrace every aspect of a patient’s identity rather than using the traditional sterile approach in technology-based care. A comprehensive list of positives, negatives and desired features was generated to guide the development of a resource to support AABN (Table 3).

**Table 3.** Thematic results for the use of technology.

| **Table 3. Thematic results for the use of technology.** |  |
| --- | --- |
| **Positives** | **Negatives** |
| · First place to look for answers, before a physician<br>· Convenient way to get information<br>· Keeps people connected<br>· Fascinating<br>· Necessary | · Can be intimidating<br>· Caution needed about information sources<br>· Can be overused<br>· Distracting |
| **Desired Features** |  |
| **Meaningful Connections**<br>· Ability to FaceTime a professional, preferably the same person throughout pregnancy and postpartum periods<br>· A tailored approach that considers “me, my history” not just references “studies, books, and other patients”<br>· Connection points: message boards that let you talk to people going through the same thing<br>**Trust and Respect**<br>· Honest, realistic feedback and unbiased opinions<br>· Culturally relevant advice and homeopathic treatments<br>**Multiple Sources of Information**<br>· The ability to cross-reference sources and form an educated opinion independently<br>· Visual tutorials on pregnancy care, how to breastfeed, and practices for overall wellness<br>· A format like WebMD: stories that are relevant right now, ability to find a doctor, and check symptoms |  |

### Representative Quotes

#### Meaningful Connections

Participants emphasized the importance of genuine connections with healthcare experts and other birthing people. FaceTime, message boards, and opportunities to chat were all accepted forms of communication. There was strong enthusiasm around finding healthcare experts who take the time to listen to each patient’s unique situation. Listing off risk factors without listening to patient backgrounds and concerns was described as off-putting by multiple focus group participants.

> *“You know, you want to go to the doctor and they’re real and they know you, your history. Don’t tell me about studies, no books, no other patients because that’s not me. I had a book doctor that could quote anything off a book and I don’t wanna hear that. You’re not me, we’re talking about me, my history, my records, and if you really haven’t been with me a long time, I’m definitely not gonna listen to you.”*
>
> *- Participant C, Focus Group 1*

> *“We know the internet can be a sketchy place, but doctors are so disconnected from the everyday life of what it takes, and what you’re going through, and they think they know everything. My situation is different, it’s not the same as hers, I don’t care what the risk factors look like. Don’t put me in a box, look at different experiences and tell me how it might be helpful to my situation.”*
>
> *- Participant L, Focus Group 2*

> *“On [pregnancy resource], a lot of moms are going through the same conditions, have the same symptoms. When you see so many people going through the same situation that you’re going through and they try different things that the doctor didn’t tell you to do and it seems relatively safe, and then it works. That’s the power of the community on the internet*.*”*
>
> *- Participant R, Focus Group 3*

> *“Having someone in person that you can speak to about your medical history, about your concerns in real time is very beneficial, and it doesn’t matter if that person is face-to-face or virtual. On the internet, if there is an option to have a virtual chat, I will utilize those because I can say, hey this is what I’m dealing with. I know what your website says this, but this wasn’t covered. That would make me more likely to use technology and trust it.”*
>
> *Participant P, Focus Group 3*

##### Trust and Shared Values

Regarding breastfeeding especially, women shared frustration with peers who only presented the benefits and connectedness, without realistic conversations about side effects or downsides. A resource was desired where women could share the benefits, but also talk realistically about pain, supply and storage issues, or building support. In addition, women wanted to speak with people who shared and respected their values. This was mentioned as an advantage to internet-based connections since there is time to research a health professional and determine if this professional’s approach to health mirrors their own.

> *“Give me the whole picture. It makes me feel good because you’re not sugarcoating it just for the sale, you know what I’m saying? Like if you’re trying to sell some juice, and you’re not saying it’s the best thing ever made. Tell me it’s bitter, but it’s going to benefit me, so take it with something*.*”*
>
> *- Participant E, Focus Group 1*

> *“With breastfeeding they always tell you the pros and not the cons. I feel like they tell you it’s safe but what about the bad stuff? There’s always side effects to everything. Always pros and cons. Tell me everything. Let me decide*.*”*
>
> *- Participant H, Focus Group 2*

> *“If I’m online, I know that I can go to places where people hold my same values. I’m much more likely to trust someone on the internet vs. in person, because I know they’re knowledgeable about what I’m looking for. I can control where I’m getting my internet stuff from. I can’t always control who the doctor in front of me is going to be*.*”*
>
> *- Participant Q, Focus Group 3*

#### Multiple Sources of Information

Participants were not interested in resources that were text alone, or only contained one source of information. Of those who enjoyed reading health-related resources, they preferred formats that were engaging, culturally relevant, and didn’t push pharmaceuticals. Other sources of information preferred by participants included videos, live classes that could be viewed online, and Q&A sessions with experts in both clinical and homeopathic medicine.

> *“If you just give something to read, I’m not going to do it. I need something I can see, I’m a visual learner. I can read something all day, 50-60 times and when I put it down, I forget what it says. If I can see what it’s doing, I remember*.*”*
>
> *- Participant A, Focus Group 1*

> *“I love to read but it’s gotta be like urban fiction, it’s gotta be something that really catches my eye and I’m not putting it down. When my baby has a fever, I go to WebMD. I gotta go somewhere where some doctor sites some reliable source. Can’t go to no Wikipedia, I’m like, nah, I’m good. I find stuff on there like*…*this don’t sound right*.*”*
>
> *- Participant B, Focus Group 1*

> *“I know doctors are there for a reason, they have some schooling, but I feel like my experience in my family, they’ve not always given the best advice, because, you know, pharmaceutical companies they’re just the richest companies in the world. I would rather look things up, cross reference them, find books, and check if articles are right rather than just believing one doctor*.*”*
>
> *- Participant J, Focus Group 2*

## DISCUSSION

Survey and focus group results highlight the feasibility of using technology to expand the African American Breastfeeding Network’s reach and the key features required to make a technology-based intervention successful. Interestingly, of the survey responses, the response with the highest, or “most likely” response was that participants were likely to look for health-related information on phones or the internet (4.23 ± 1.00, between “Likely” and “Very Likely”), however, the lowest, or “least likely” response was regarding the likelihood of trusting that information (3.04 ± 0.80, or “Somewhat Likely”). Focus groups outlined the positives and negatives of technology usage and identified that a resource would need to: 1) build meaningful connections, 2) build a community based on trust and shared values, and 3) include multiple sources of information in different formats, to appeal to different kinds of learners.

Community-driven and culturally aware health-related technology has great potential to build stronger bridges between patients and desired healthcare services. In one study, researchers revealed that when considering the entire Black patient population, an overall disparity exists between Black and NHW patients regarding access to telemedicine. However, in younger, female Black patient populations, there is an increased uptake of telehealth resources.^11^ Black, Latino, and Filipino seniors are likely to have less access to digital tools, less experience performing a variety of online tasks, and are less likely to feel capable of going online for health information, however, this is not representative of all age groups.^12^ Our findings supported this study, with all Black women participating in our focus group demonstrating some willingness to engage with telehealth resources.

We found one other study about mobile health technology specific to breastfeeding support. In this study, fifty pregnant and postpartum African Americans from the District of Columbia identified a preference for apps with: 1) text messaging technology, 2) social support and desirability, and 3) support for topics like parent-child attachment and returning to work. Like our focus group participants, women in this study asked for peer discussions as a feature of mobile health technology.^13^ In a study looking at technology and COVID-19 communications, investigators found that Black respondents were more likely than NHW respondents to report posting COVID-19-related content on social media.^14^ Understanding technological challenges and engagement in populations of interest is critical to ensure that technology is utilized to improve healthcare for all.

Initially, our research team intended to use this information to inform the development of an app or website that would expand AABN’s reach. However, with the onset of the COVID-19 pandemic occurring shortly after the data was analyzed, the information was instead used to apply for funding mechanisms that would transition some birthing support services from in-person to virtual settings. As a result, since 2020, the WeRise doulas have worked in Southeastern Wisconsin to provide continuous physical, emotional, and informational support to birthing persons before, during, and shortly after childbirth to help them achieve the healthiest, most satisfying experience possible.^15^

### Meaningful Connections

In a study detailing the impact of community-based doulas on low-income Black birthing families, positive impacts were credited to similarities in race and culture, support and resources beyond birth, and the ability of a community-based doula to recognize the institutional biases that exist in the health care system and mediate their effect on birthing persons.^16^ In an interview with a pregnant mother of two, who described her last two birth experiences as traumatic, WeRise doulas were credited with helping her find a voice and express her needs and wants.^17^ During COVID-19, in addition to virtual consultations, WeRise doulas provided masked, in person support, and AABN hosted outdoor community gatherings to hand out supplies and care packages for birthing families.

### Trust and Shared Values

WeRise doulas leverage training and experience to educate Black birthing families, understand wants and needs, and draw up birth plans. Because of this time spent building relationships and connecting on the most important aspects of labor and delivery, the doula can advocate for the client’s wishes and help prevent traumatic birth experiences. In one situation with a WeRise doula, a client had to be rushed into an emergency C-section, and the doula had a backup plan ready, switching to practicing affirmations with the client to help her feel calm and empowered.^18^

### Multiple Sources of Information

As community-based doulas in the WeRise initiative work to support Black birthing families, they face similar challenges to those across the country. Other studies list COVID-19-related fear of exposure, limited access to support systems, and uncertainties surrounding hospital restrictions on labor and birth.^19^ Whether information is delivered via technology or in-person, equitable integration into the clinical care team is vital to ensure that the birthing family receives multiple sources of accessible information and selects options that are best for their birthing experience.^18,19^

## CONCLUSION

Overall, the demand for AABN services remains high, and there is an urgent need to create sustainable tools to support birthing services. Moving toward digital health equity requires a community-based and community-driven approach. Future studies will disseminate specific, tailored methods that AABN birth workers use to support birthing families, through technology-based platforms, and measure their impact on maternal and child health outcomes.

## Data Availability

All data produced in the present study are available upon reasonable request to the authors.

## Notes

### Competing Interest Statement

The authors have declared no competing interest.

### Funding Statement

This study did not receive any funding.

### Author Declarations

All data collection and research-related actions were completed under approval from the Medical College of Wisconsins Institutional Review Board (PRO#00032076).

